# Glymphatic Flow after Thrombectomy is Associated with Futile Recanalization in Large Vessel Occlusion Stroke

**DOI:** 10.1101/2025.01.16.25320702

**Authors:** Alan J. Finkelstein, Matthew T. Sipple, Sajal M. Akkipeddi, Racquel White, Gurkirat Singh Kohli, Stephen Susa, Rohin Singh, Prasanth Romiyo, Jianhui Zhong, Tarun Bhalla, Thomas Mattingly, Vincent Nguyen, Maiken Nedergaard, Matthew T. Bender, Derrek Schartz

**Affiliations:** University of Rochester, Department of Biomedical Engineering; University of Rochester School of Medicine & Dentistry; University of Rochestery, Department of Neurological Surgery; University of Rochester, Department of Imaging Sciences; University of Rochester, Department of Neurology

**Keywords:** Ischemic Stroke, Futile Recanalization, Glymphatic System, DTI-ALPS, Diffusion Imaging, Thrombectomy

## Abstract

**Background:** Stroke is a leading cause of global death and disability, with mechanical thrombectomy remaining the optimal treatment approach for large vessel occlusion (LVO) ischemic stroke. Despite endovascular recanalization, nearly half of patients experience poor functional outcomes, a phenomenon termed “futile recanalization”. The cerebral glymphatic system has emerged as a potential, yet underexplored therapeutic target. In this retrospective study, we utilized glymphatic diffusion tensor analysis on post-thrombectomy MRIs scans to evaluate the association between glymphatic flow, clinical outcomes, and futile recanalization in LVO ischemic stroke.

**Methods:** In this retrospective study, 133 patients with anterior LVO ischemic stroke and MRI post-thrombectomy were identified. Futile recanalization was defined as a modified Rankin score >2 at 90 days, despite achieving complete or near-complete angiographic recanalization (mTICI 2b-3). We employed diffusion tensor imaging along the perivascular space (DTI-ALPS) to evaluate glymphatic function in individuals with futile recanalization and those with functional independence at 90 days. Spearman’s rank correlation was used to examine associations between the ALPS index and clinical variables. Effect sizes were calculated and reported using Cohen’s d.

**Results:** In total, 55 anterior circulation LVO patients (mean age 73.9, 44% male) were included with adequate post-thrombectomy MRIs for analysis. Overall, glymphatic clearance was lower in the infarct side compared to the contralateral side (p = 0.035). Patients with futile recanalization had lower glymphatic flow compared to those with functional independence (p=0.049). Additionally, glymphatic flow was significantly associated with presenting NIHSS (R=-0.46, p = 0.002).

**Conclusions:** These findings suggest that patients with futile recanalization have comparatively worse glymphatic clearance. Further research is required to clarify the relationship between futile recanalization and the glymphatic system, which may facilitate the development of therapeutic adjuncts.

## INTRODUCTION

Stroke is a leading cause of global death and disability, with mechanical thrombectomy being the optimal treatment approach for large vessel occlusion (LVO) ischemic stroke^1^. However, despite successful endovascular recanalization of the culprit LVO, approximately half of patients still have poor functional outcomes, which is referred to as “futile recanalization”^2^. The complex pathophysiology of futile recanalization is thought to involve arterial reocclusion, poor collateral circulation, incomplete microcirculatory reperfusion, and disruption of the blood-brain barrier (BBB)^3,4^. As a result, identifying potential therapeutic targets for developing adjunctive brain cytoprotective therapies following recanalization is of utmost importance to further maximize the benefit of endovascular therapy^3,5^. One understudied and potential therapeutic target is the cerebral glymphatic system.

The glymphatic system comprises perivascular channels formed by astroglial cells to promote the elimination of proteins, distribute metabolites, and maintain homeostasis of cerebrospinal fluid (CSF) ^6^. CSF flows along perivascular spaces into the brain parenchyma mediated by aquaporin 4 (AQP4) and arterial pulsation. This results in the flow of CSF from the subarachnoid space, enabling the exchange of CSF and interstitial fluid (ISF). Ultimately, solutes are drained into the venous perivascular spaces and meningeal lymphatics. Dysfunction of the glymphatic system has been implicated in many diseases, including Alzheimer’s Disease, cerebral small vessel disease (CSVD), and stroke, due to impaired waste removal, autonomic dysregulation, and diminished cerebral perfusion^7^.

Autonomic dysregulation is observed following ischemia^8^, thereby modulating arterial pulsation and glymphatic function. Following acute ischemic injury, glymphatic dysfunction also contributes to the formation of brain edema, mediated by a rapid perivascular influx of CSF into the brain parenchyma^9,10^. Further, pan-adrenergic blockade has been shown to potentiate glymphatic clearance, normalizing cerebral edema^11^. Relatedly, functional hyperemia, or neurovascular coupling, results in increased arterial pulsatility, thereby improving glymphatic function and removal of metabolic waste^12^. Thus, potentiating glymphatic drainage may ameliorate post-ischemic and traumatic edema^13^ as well as remove accumulated cytotoxic solutes. However, it remains largely unknown how glymphatic clearance after mechanical thrombectomy is associated with clinical outcomes in LVO ischemic stroke.

Advances in neuroimaging enable non-invasive investigation of the cerebrum. Diffusion-weighted imaging (DWI) is a method to probe water movement within the brain parenchyma^14^. Diffusion tensor imaging along the perivascular space (DTI-ALPS) is an advanced DWI method devised to evaluate the glymphatic system^15^non-invasively. This technique measures diffuson in the radial direction at the level of the lateral ventricular body adjacent to the perivascular spaces of the deep medullary draining veins, which is a primary drainage pathway of the glymphatic system. The DTI-ALPS method has demonstrated robustness^16^ and has been used to investigate impaired glymphatic clearance in various neuropathologies^15,17,18^. In this retrospective study, we applied DTI-ALPS on post-thrombectomy MRIs to investigate if glymphatic flow is associated with clinical outcomes and futile recanalization in LVO ischemic stroke.

## METHODS

### Patient Cohort

This study was approved by the University of Rochester Medical Center (URMC) Research Subject Review Board. Need for patient consent was waived by the URMC institutional review board. Stroke patients who were treated with mechanical thrombectomy from 2017 to 2021 were retrospectively reviewed from a prospectively maintained database from our single-center tertiary healthcare system. All mechanical thrombectomies were performed within 24 hours from symptom onset and did not include large-core infarcts. Patients who underwent a post-thrombectomy MRI with sufficient imaging for analysis (further detailed below) were included within the study. A brain MRI is typically acquired at our institution as part of standard-of-care and is usually obtained 24 hours after thrombectomy. To reduce potential confounding variables related to occlusion location (e.g. large vessel vs. medium vessel; anterior circulation vs. posterior circulation), only large vessel anterior circulation occlusions involving the internal carotid artery (ICA) and/or M1 segment of the middle cerebral artery (MCA) were included in the presented analysis. Numerous patient, clinical, and procedural variables were collected and presented. Infarct volume on MRI was determined using the ABC/2 method as previously described^19^. Futile recanalization was defined as poor functional outcomes (that is, an mRS >2 at 90 days) despite having achieved complete or near-complete angiographic recanalization (mTICI 2b-3) via endovascular thrombectomy. ‘Functionally Independent’ was defined as achieving functional independence (mRS 0-2) at 90 days after endovascular recanalization. The STROBE corss sectional checklist was used when writing our report^20^.

### Magnetic Resonance Image Acquisition

Post-thrombectomy patient brain MRIs were obtained using a 3T GE DISCOVERY scanner (General Electric, Waukesha, WI, USA) with a 32-channel head coil, usually approximately 24 hours after intervention. 2D axial diffusion-weighted imaging was acquired using a spin-echo echo planar imaging with PROPELLER (SE-EPI) technique, with 12 diffusion directions (b=1000 s/mm^2^) and 2 b=0 s/mm^2^ reference images. Scan parameters were: TR/TE = 1000/79.7 ms, echo spacing = 0.18 ms, voxel size = 1.0 × 1.0 × 3.6 mm³, slice thickness = 3.6 mm, matrix size = 256×128, and a field-of-view (FOV) of 256×256 mm^2^, with SENSE in-plane acceleration R=2. For 2D axial T1-weighted imaging, a spin-echo sequence with PROPELLER was used (TR/TE = 2300/50 ms, echo train length (ETL) = 16, TI = 950 ms, flip angle = 111°, voxel size = 0.5 × 0.5 × 6 mm³, slice thickness = 5 mm, FOV = 512×512 mm^2^, matrix size = 512×288 mm^2^), also with SENSE in-plane acceleration R=2.

### Glymphatic Diffusion Tensor Imaging Along Perivascular Space (DTI-ALPS) Analysis

The diffusion tensor imaging along the perivascular space (DTI-ALPS) method was used to measure the glymphatic flow of the post-thrombectomy MRIs^15^. This method has been extensively used and described in prior studies^15,17,21,22^. The ALPS index, a surrogate measure of glymphatic flow, is calculated by evaluating water diffusivity along projection and association fibers adjacent to the lateral ventricle body, which can be measured using standard regions of interest (ROIs)^22^(Figure 1). Preprocessing was performed using MRtrix3^23^ including global intensity normalization and denoising. Subsequently, denoised data were corrected for eddy current-induced distortion using EDDY_CORRECT in FSL and addressed susceptibility-induced distortion using INVERSION^24^, consisting of inverse contrast normalization of T1w data and diffeomorphic co-registration using symmetric normalization in advanced normalization tools (ANTs)^25^. Diffusivity maps were computed along the x-axis (Dxx), y-axis (Dyy), and z-axis (Dzz), along with fractional anisotropy (FA), mean diffusivity (MD), axial diffusivity (AD), and radial diffusivity (RD) using DIPY. FA maps were co-registered to the FA map template FMRIB58 atlas using ANTs. The FA registration matrix was used to warp all other DTI maps to standard space. Modified Johns Hopkins University ICBM white matter labels were used for the projection (superior corona radiata) and association fibers (superior longitudinal fasciculus) in the periventricular area, as previously described^17,22^. For quality assurance purposes, ROIs were defined in standard (MNI) space (or, FMRIB58), and all diffusion directions were warped to MNI/FMRIB58 space. Infarcts that involved the ROI were excluded from analysis. An example region of interest determination in a post-thrombectomy patient can be seen in **Figure 1**. The ALPS index was calculated using these labels and defined by:

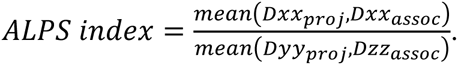

**Figure 1.**
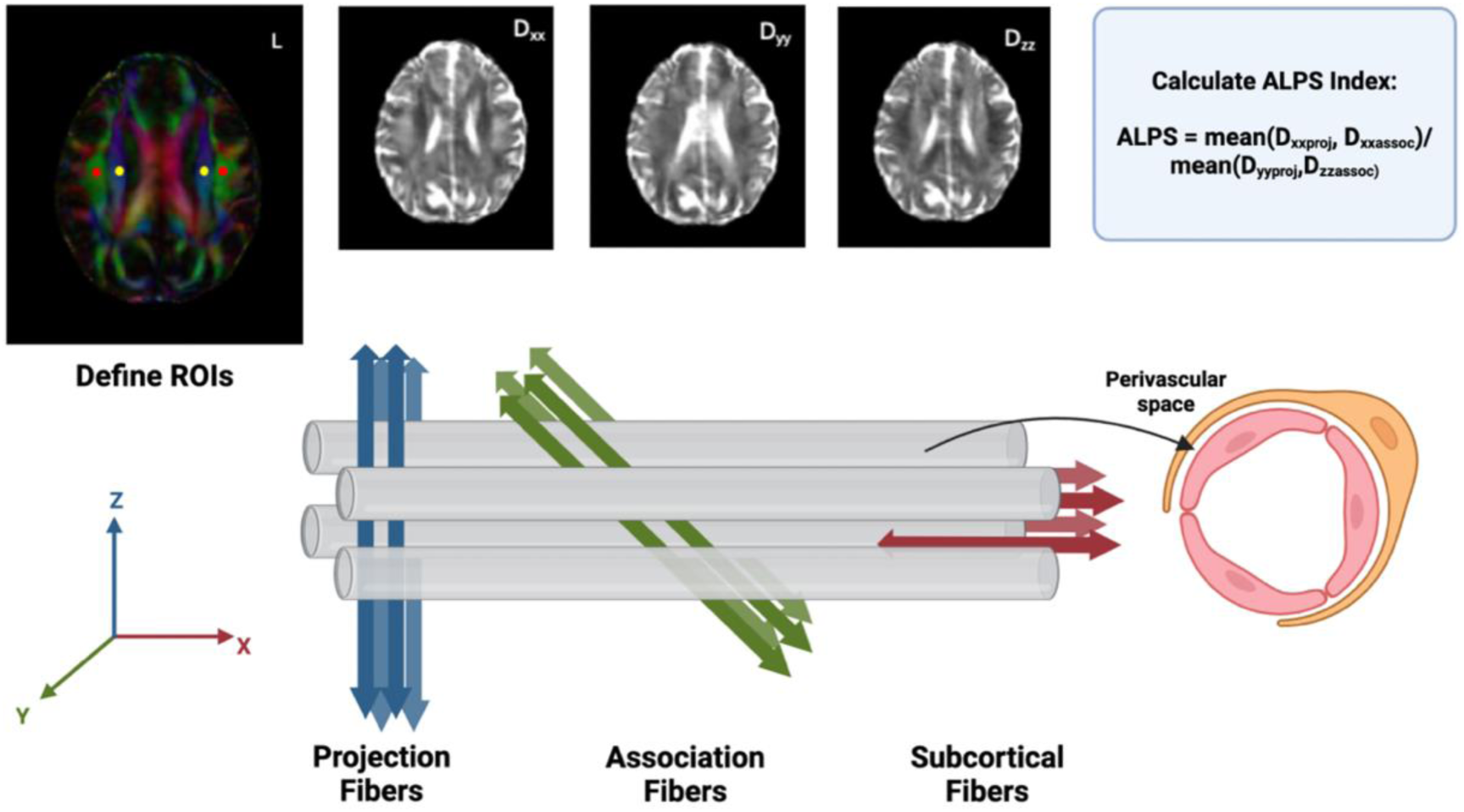
Overview of DTI-ALPS principle. Diffusion-weighted data is acquired along 12 non-collinear directions from which diffusion-tensor analysis is performed to estimate the diffusion tensor. Regions of interest are defined in standard space. Diffusion along the x, y, and z direction are estimated from the diagonals of the diffusion tensor and used to calculate the diffusion along the x, y, and z directions within each ROI. The blue, green, and red regions correspond to projection fibers, association fibers, and subcortical fibers, respectively. The ALPS-index is then calculated accordingly which corresponds to flow within the perivascular space.

The ALPS-index was calculated for both the infarct and non-infarct side. The mean ALPS-index was calculated as the average between the ALPS-index for the infarcted and non-infarcted side.

### Statistical Analysis

Exploratory data analyses were performed using a heatmap using the non-parametric Spearman rho correlation coefficient to evaluate univariate associations between variables. Paired t-tests were conducted to compare outcomes between paired data. The magnitude of the differences between groups was assessed by calculating effect sizes using Cohen’s d. Non-parametric Spearman’s rank correlation coefficient was used to examine univariate associations between neuroimaging and clinical variables. Differences between categorical variables were evaluated using an analysis of covariance (ANCOVA) with sex and age included as covariates. Chi-square test of independence was conducted to examine the difference of rates between categorical variables. Adjusted p-values were obtained using the Benjamini-Hochberg method. For all statistical analyses, an adjusted p-value <0.05 was considered significant. All statistical analyses were performed in R (R version 4.1.1, Vienna, Austria; https://www.r-project.org/).

## RESULTS

### Study Population and Baseline Patient Characteristics

In total, 329 patients were identified during the study period who had underwent a mechanical thrombectomy for acute ischemic stroke. From these, 193 (58.7%) had an isolated anterior circulation LVO involving the ICA and/or M1 division of the MCA. Eighty-six (44.6%) of these were excluded since they either did not have a post-thrombectomy MRI or did not have sufficient sequences for analysis. Thus, 107 LVO patients had sufficient MRIs for potential inclusion. However, 52 (48.6%) were excluded if the infarct involved the ROI required for glymphatic DTI analysis with evidence of mass effect. As a result, 55 (28.5%) anterior circulation LVO patients with sufficient imaging were included in the final analysis (**Figure 2**).

**Figure 2.**
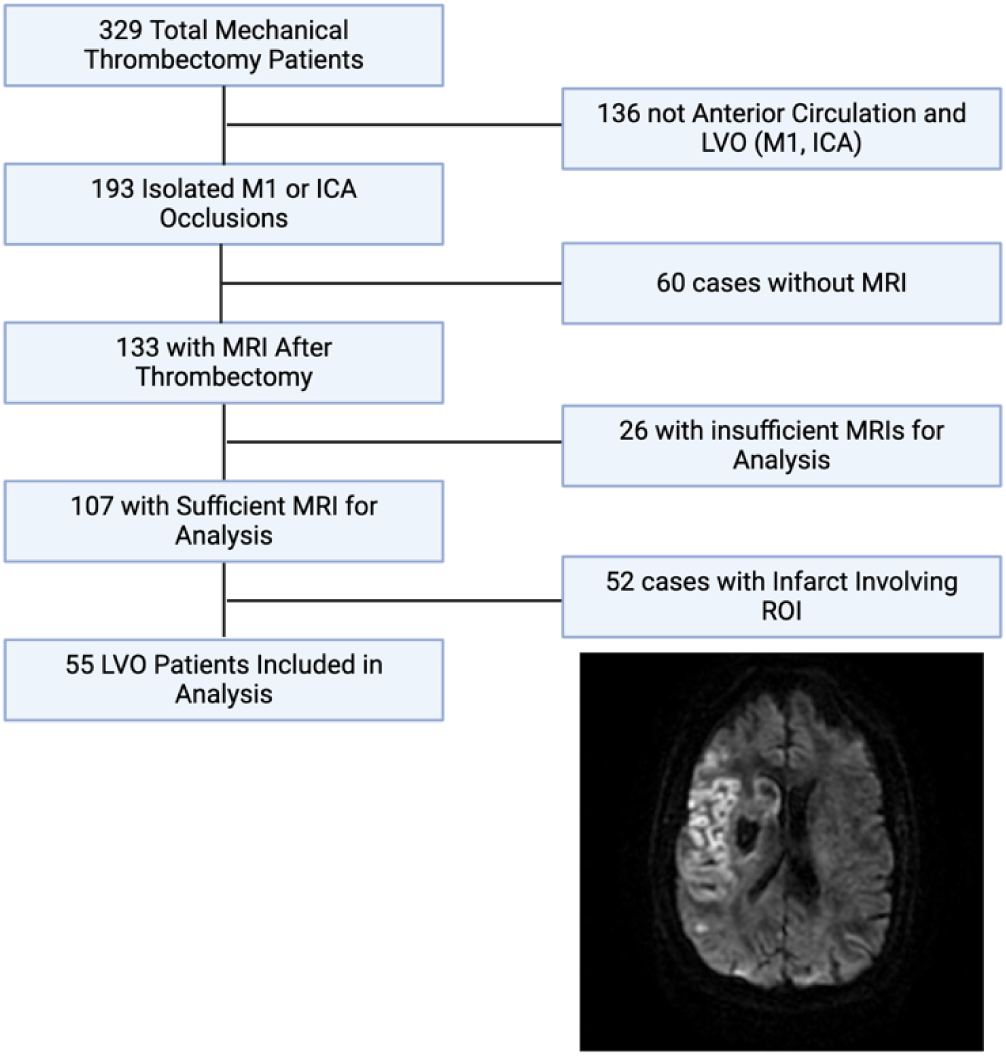
(A) Overview of inclusion/exclusion criteria for selection of final study population, along with an example of an excluded patient due to involvement of the basal ganglia and mass effect. (B) Example of an individual with futile recanalization and functional independence.

Of the 55 included patients, the average age was 73.9 (SD: 13.2) years old and 44% (24/55) were male. Occlusion location was the M1 segment of the MCA in 44/56 (78.6%) and the terminal ICA in 11/55 (20%) of cases. Post-thrombectomy MRI was most often acquired at approximately 24 hours after intervention (41/55, 74.5%), followed by 48 hours (8/55, 14.5%), and 72 or 12 hours (both 3/55, 5.5%). The mean presenting ASPECT Score was 8.8 (SD: 1.1) and NIHSS was 15.7 (SD: 7.1). Successful endovascular final recanalization (mTICI 2b-3) was achieved in 96% (53/55) of patients. Futile recanalization occurred in 21/53 (40%) of these patients. For full baseline patient characteristics, refer to **Table 1**.

**Table 1:** Overall baselines characteristics and demographics of cohort.

| Variable | N (%) |
| --- | --- |
| *Patient Demographics* |  |
| Total Patients | 55 (100) |
| Mean Age | 73.9 (SD: 13.2) |
| Male | 24 (44) |
| White | 44 (80) |
| Black | 6 (11) |
| Other ethnicity | 5 (9) |
| *Past Medical History* |  |
| Atrial Fibrillation | 25 (45) |
| Diabetes Mellitus | 16 (29) |
| Coronary Artery Disease | 13 (24) |
| Cancer, Active | 6 (11) |
| Hyperlipidemia | 26 (47) |
| Hypertension | 38 (69) |
| Smoking, Current | 8 (15) |
| Prior Stroke | 8 (15) |
| *Presentation and Treatment* |  |
| M1 Occlusion | 44 (80) |
| ICA Occlusion | 11 (20) |
| Mean NIHSS | 15.7 (SD: 7.1) |
| Mean ASPECTS | 8.8 (SD: 1.1) |
| Intravenous tPA | 27 (49) |
| Mean LNW to Recanalization (min) | 417 (SD: 362) |
| *Patient Outcomes* |  |
| Successful Recanalization (mTICI 2b-3) | 53 (96) |
| 90-day mRS 0-1 | 25 (45) |
| 90-day mRS 0-2 | 33 (60) |
| 90-day mortality | 8 (15) |
| Futile Recanalization | 21 (40) |

Cohort specific statistics are enumerated in **Table 2** comparing the differences between patients with futile recanalization and those with functional independence at 90 days. No differences between groups were observed between timing of post op MRI (p=0.259), time to recanalization (p = 0.918), sex (p-=0.99), or the ASPECTS score (p=0.749). However, age, glymphatic flow (mean ALPS-index), and infarct volume were significant different between the two groups (p<0.05).

**Table 2:** Comparisons of specific variables between individuals with futile recanalization and functional independence. ASPECTS : Alberta stroke program early CT score; ALPS: Diffusion tensor imaging along the perivascular space. M: Male, F: Female. A p-value < 0.05 is considered significant.

| Variable | Futile Recanalization | Functional Independence | P-value | Cohen's d |
| --- | --- | --- | --- | --- |
| Sex [M/F] | 9/12 | 15/17 | 0.99 | -0.0792 |
| Age (mean [sd] ) | 81.1 [8.47] | 68.8 [13.9] | **0.0002** | -1.07 |
| Mean ALPS-Index | 0.974[0.0667] | 1.01[0.0630] | **0.0493** | 0.572 |
| Infarct Volume (mean [sd] in cm<sup>3</sup>) | 57.4 [64.0] | 24.0 [36.8] | **0.0385** | -0.640 |
| ASPECTS | 8.71 [1.10] | 8.08 [1.06] | 0.749 | 0.0908 |
| Onset time to Recanalization (mean [sd] in minutes) | 428 [394 ] | 417 [356 ] | 0.918 | -0.0293 |
| Timing of Post Op MRI (mean [sd] in hours) | 32.6 [16.6] | 27.8 [12.0 ] | 0.259 | -0.333 |

### Post-Thrombectomy Diffusion Tensor Glymphatic Results

On post-thrombectomy MRI, the ALPS-index was significantly lower on the infarct side compared to the non-infarct side (0.982 [SD: 0.082] vs 1.02 [SD: 0.084] , respectively, p= 0.035), with a moderate effect size (Cohen’s d = -0.33; Figure 3A). Figure 3b displays a heatmap illustrating univariate associations between continuous variables within the cohort. Strong significant (p<0.05) positive associations were observed between cerebrovascular risk factors and outcomes such as hypertension, and older age.

**Figure 3.**
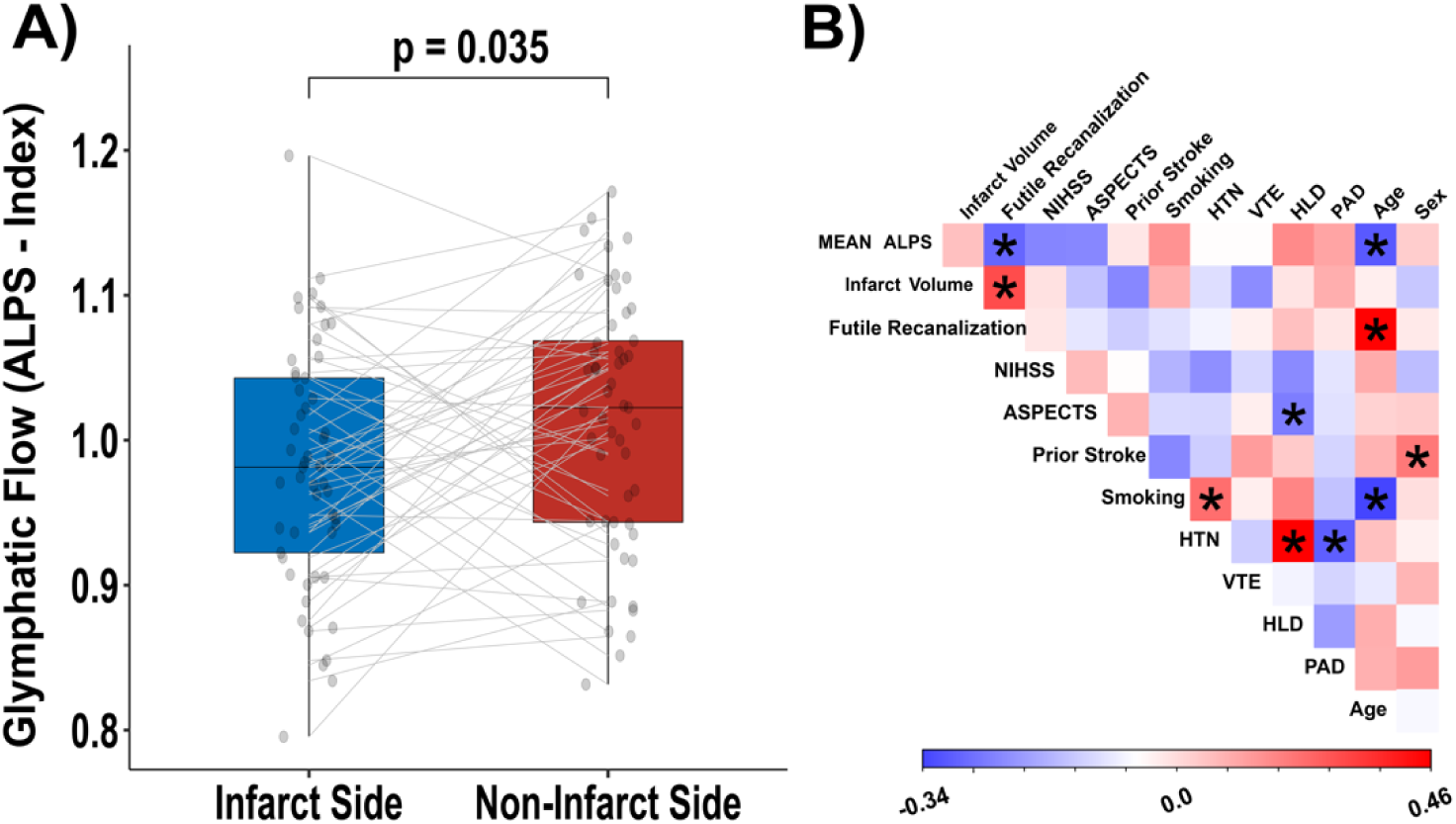
(A) Box-and-whisker plot illustrating the differences in the ALPS index between the infarct side and the contralateral, non-infarcted side. (B) Heatmap illustrating univariate assocations between continuous variables, using the non-parametric Spearman rank correlation coefficient. An asterisk indicates a statistically significant association. NIHSS = NIH stroke scale; RAPID = rapid response team ; HTN = Hypertension; VTE = Venous thromboembolism; HLD = Hyperlipidemia; PAD = peripheral arterial disease; mRS = modified Rankin Scale.

Mean glymphatic flow was significantly lower in individuals with futile recanalization (**Figure 4A**) (mean ALPS-index = 1.03 [SD: 0.059] vs 0.972 [SD: 0.060], respectively, p = 0.049) with a moderate effect size (Cohen’s d = 0.572). Furthermore, the mean ALPS-index was inversely related to the NIHSS in those with futile recanalization (R=-0.46, p = 0.00213), which was not observed in those with functional independence (R = -0.016, p=0.899; **Figure 4B**). This relationship is further emphasized by an observed interaction effect between recanalization outcomes and NIHSS (𝛽 = -0.0032, p = 0.0163), indicating the relationship between glymphatic function and NIHSS differs by recanalization status.

**Figure 4.**
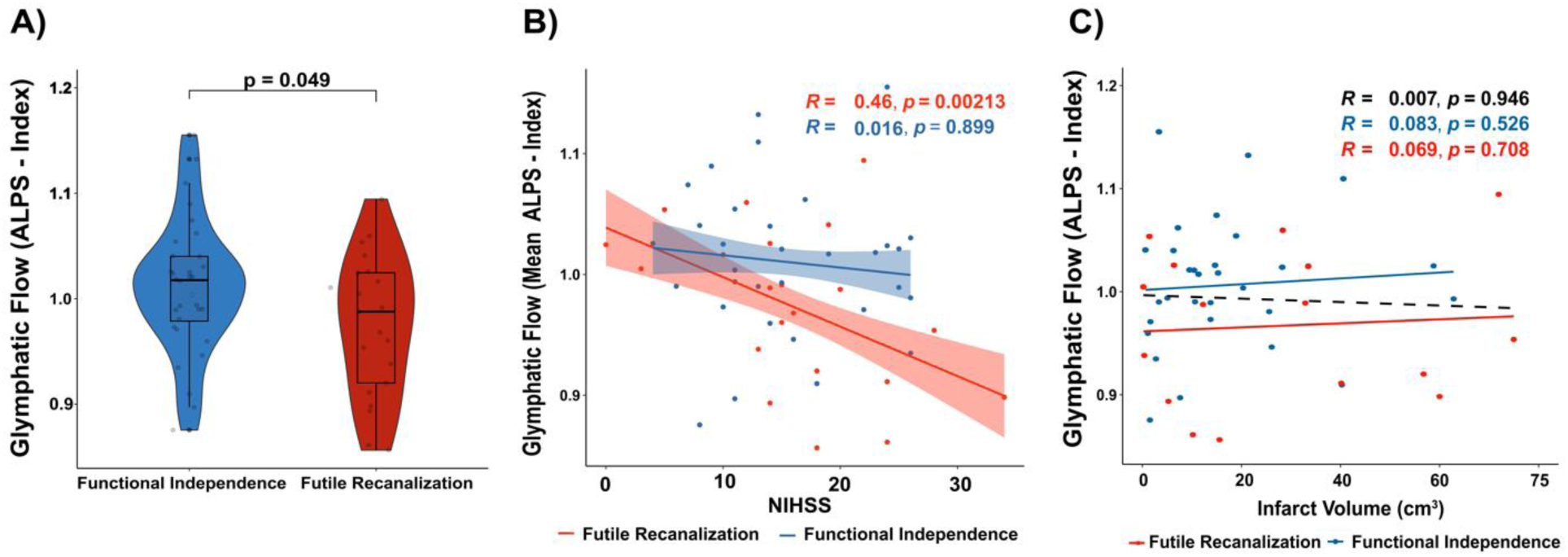
(A) Violin with Box-and-whisker plots illustrating differences in the mean ALPS index between individuals with futile recanalization and those with functional independence at 90 days. **(B)** Association between the mean ALPS index and the NIHSS stratified based on futile recanalization. Lines represent the linear regression fits for each group, with corresponding correlation coefficients and p-values annotated. Shaded regions represent the 95% confidence intervals for the best-fit line. NIHSS: NIH Stroke Scale. **(C)** Association between glymphatic flow (ALPS-index) and infarct volume stratified based on futile recanalization. Lines represent the linear regression fits for each group, with corresponding correlation coefficients and p-values annotated. The black dashed line represents the linear regression fit for the entire cohort.

Infarct volume was significantly smaller in those with functional independence compared to those with futile recanalization (mean infarct volume in cm^3^ = 23.96 [SD: 36.82] vs 57.37 [SD: 64.03], respectively, p = 0.039) with a moderate effect size (Cohen’s d = -0.647, Table 2). No association was observed between glymphatic flow and infarct volume in patients with futile recanalization (R = 0.099, p =0.565), functional independence at 90 days (R = 0.11, p = 0.413), or across the entire cohort (R = 0.00018, p=0.0986) (**Figure 4C**).

## DISCUSSION

This study leveraged non-invasive diffusion tensor imaging measurements of the glymphatic system and found that glymphatic flow was significantly lower in the infarcted hemisphere compared to the contralateral side. Furthermore, we found a direct association between post-thrombectomy glymphatic flow and futile recanalization at 90-day follow-up. Likewise, we observed a significant association between presenting stroke severity and glymphatic flow in patients with futile recanalization. Taken together, these data provide in-human evidence suggesting that alterations in the glymphatic system might play an important role in stroke presentation and functional outcomes after thrombectomy in acute LVO ischemic stroke.

Animal models and prior studies in humans have indicated that gymphatic flow is lower in the infarcted side in ischemic stroke. Prior to thrombectomy, MR indices of glymphatic flow have been shown to be lower in the infarcted side compared to the non-infarcted side^26^. A lower ALPS-index has also been shown to be correlated with deficits in motor function^27^. In the presented work, the ALPS-index of the infarcted side was similarly lower than the contralateral side following thrombectomy. Previous work has also shown a lower ALPS index in the infarcted side was associated with functional outcomes and presenting stroke severity^28^. Relatedly, ischemic stroke mouse model studies have demonstrated impaired parivascular CSF circulation, revealing glymphatic system dysfunction as a primary driver of cytotoxic brain edema^28,29^.

The presented investigation found that glymphatic impairment was associated with greater presenting stroke severity and futile recanalization. A lower ALPS-index correlated with a higher presenting NIHSS score, particularly for patients who did not achieve functional independence at 90 days (Figure 4b). We hypothesize that this might relect either the degree of ischemic insult and/or the patient’s baseline glymphatic function which may facilitate neurologic recovery. However, we did not find an association observed between glymphatic function and infarct volume, suggesting these measures might represent related but distinct pathophysiological processes. However, it has been shown that glymphatic function is mediated in part by astrocytes, which are are particularly vulnerable to early ischemic injury, resulting in cytotoxic edema^10,30^. Conversely, blood brain barrier dynsruption^31^ resulting in vasogenic edema and larger infarct volumes. As a possible explanation for our findings, we suspect that in patients with functional independence, functional hyperemia and robust microcirculatory reperfusion is achieved which facilitates improved clearance of cytotoxic solutes^12^, explaining the higher glymphatic flow in patients with good functional outcomes. Future studies are needed to elucidate the precise role of glymphatic dysfunction in futile recanalization to identify therapeutic targets for improve outcomes following mechanical thrombectomy.

Modulation of the glymphatic system may be important to improving outcomes among patients who currently experience futile recanalization following endovascular therapy. In acute cerebral injury autonomic dysregulation results in increased concentrations of catecholamines in the CSF and plasma, resulting in diminished glymphatic efflux^8,11,13,32^. Adrenergic receptor antagonism potentiates glymphatic clearance and promotes CSF exchange and normalization of brain parenchymal extracellular fluid, facilitating reduced infarct volume and improved motor outcomes in an ischemic stroke mouse model^11^. Likewise glymphatic system dysfunction in acute ischemia is a key contributor to the formation of brain edema, while, adrenergic inhibition can alleviate cytotoxic edema and reduces post-ischemia cognitive impairment^28^. Mechanistically, interstitial fluid stagnation with an accumulation of neuroinflammatory and cellular debris likely results in decreased glymphatic function in patients with futile recanalization^11,13^. Thus, it is possible pan-adrenergic blockage could function as a therapeutic adjunct to endovascular thrombectomy, which should be explored in future studies.

In addition to adrenergic receptors, aquaporins represent a potential therapeutic target for glymphatic system modulation in large vessel occlusion ischemic stroke. Dysfunction of tight-junctions and endothelial barrier results in BBB dysfunction with disruption of pial arteries and pericytes. Thus, modulation of tight-junction proteins may also facilitate improved glymphatic function^33^. In mouse models of cytotic edema, reflective of cerebral ischemia, AQP4 knockout mice have improved clinical outcomes^34,35^. Accordingly, inhibition of AQP4 channels might also modulate glymphatic clearance and improve cerebral edema following endovascular treatment^36^. Future studies are needed to investigate the safety and efficacy of therapeutic interventions in modulating glymphatic function to improve outcomes in ischemic stroke.

This investigation has several limitations to consider. First, this was a single center retrospective analysis of 55 anterior circulation LVO patients. We included only anterior circulation occlusions of the M1 MCA and/or ICA to reduce possible confounding related to stroke location. Accordingly, the generalizability of our results to more distal anterior and posterior circulation occlusions cannot be assumed. Additionally, our sample size was relatively modest (N=55) due to the inclusion criteria of MRIs needed for analysis.Another limitation is that the infarct cannot involve the ROIs positioned within the projection and association fibers lateral to the ventricle body for this analysis pipeline. Lastly, while the ALPS-index may serve as a functional biomarker, it is not a direct measure of the glymphatic system. Rather it is an indirect measure based on the radial diffusivity within the area of presumed glymphatic drainage^37^, and may also be affected by surrounding pathology which is expected to suppress glymphatic flow^38^. There is also concern that CSF-ISF exchange does not significantly occur in the deep gray and white matter, where the ALPS-index is measured^39^. Future studies utilizing the ‘gold standard’ of intrathecal contrast-enhanced MRI^40^ might be used to verify the presented findings. Furthermore, development of novel in vivo quantitative imaging technologies will help to elucidate the pathophysiology of futile recanalization, motivating improved interventions and treatments^36,40,41^.

## CONCLUSIONS

Diffusion tensor indices of glymphatic flow are significantly decreased within the infarcted hemisphere after mechanical thrombectomy. Moreover, patients with poor functional outcomes and futile recanalization after anterior circulation LVO thrombectomy have lower glymphatic flow compared to patients with functional independence. Potentiating glymphatic clearance may function as a possible therapeutic adjunct to endovascular therapy. Larger and prospectively designed studies are required to validate these findings.

## Data Availability

The data that support the findings of this study are available from the corresponding author upon reasonable request.

## REFERENCES

1. Goyal M, Menon BK, van Zwam WH, Dippel DW, Mitchell PJ, Demchuk AM, Davalos A, Majoie CB, van der Lugt A, de Miquel MA, et al. Endovascular thrombectomy after large-vessel ischaemic stroke: a meta-analysis of individual patient data from five randomised trials. Lancet. 2016;387:1723–1731. doi: 10.1016/S0140-6736(16)00163-X

2. Shahid AH, Abbasi M, Larco JLA, Madhani SI, Liu Y, Brinjikji W, Savastano LE. Risk factors of futile recanalization following endovascular treatment in patients with large-vessel occlusion: systematic review and meta-analysis. Stroke: Vascular and Interventional Neurology. 2022;2:e000257.

3. Deng G, Chu YH, Xiao J, Shang K, Zhou LQ, Qin C, Tian DS. Risk Factors, Pathophysiologic Mechanisms, and Potential Treatment Strategies of Futile Recanalization after Endovascular Therapy in Acute Ischemic Stroke. Aging Dis. 2023;14:2096–2112. doi: 10.14336/AD.2023.0321-1

4. Luby M, Hsia AW, Nadareishvili Z, Cullison K, Pednekar N, Adil MM, Latour LL. Frequency of blood-brain barrier disruption post-endovascular therapy and multiple thrombectomy passes in acute ischemic stroke patients. Stroke. 2019;50:2241–2244.

5. El Amki M, Wegener S. Improving Cerebral Blood Flow after Arterial Recanalization: A Novel Therapeutic Strategy in Stroke. Int J Mol Sci. 2017;18. doi: 10.3390/ijms18122669

6. Jessen NA, Munk AS, Lundgaard I, Nedergaard M. The Glymphatic System: A Beginner’s Guide. Neurochem Res. 2015;40:2583–2599. doi: 10.1007/s11064-015-1581-6

7. Rasmussen MK, Mestre H, Nedergaard M. The glymphatic pathway in neurological disorders. Lancet Neurol. 2018;17:1016–1024. doi: 10.1016/S1474-4422(18)30318-1

8. Dorrance AM, Fink G. Effects of stroke on the autonomic nervous system. Compr Physiol. 2015;5:1241–1263.

9. Mestre H, Du T, Sweeney AM, Liu G, Samson AJ, Peng W, Mortensen KN, Staeger FF, Bork PAR, Bashford L, et al. Cerebrospinal fluid influx drives acute ischemic tissue swelling. Science. 2020;367. doi: 10.1126/science.aax7171

10. Ji C, Yu X, Xu W, Lenahan C, Tu S, Shao A. The role of glymphatic system in the cerebral edema formation after ischemic stroke. Experimental Neurology. 2021;340:113685.

11. Monai H, Wang X, Yahagi K, Lou N, Mestre H, Xu Q, Abe Y, Yasui M, Iwai Y, Nedergaard M, et al. Adrenergic receptor antagonism induces neuroprotection and facilitates recovery from acute ischemic stroke. Proc Natl Acad Sci U S A. 2019;116:11010–11019. doi: 10.1073/pnas.1817347116

12. Holstein-Ronsbo S, Gan Y, Giannetto MJ, Rasmussen MK, Sigurdsson B, Beinlich FRM, Rose L, Untiet V, Hablitz LM, Kelley DH, et al. Glymphatic influx and clearance are accelerated by neurovascular coupling. Nat Neurosci. 2023;26:1042–1053. doi: 10.1038/s41593-023-01327-2

13. Hussain R, Tithof J, Wang W, Cheetham-West A, Song W, Peng W, Sigurdsson B, Kim D, Sun Q, Peng S, et al. Potentiating glymphatic drainage minimizes post-traumatic cerebral oedema. Nature. 2023;623:992–1000. doi: 10.1038/s41586-023-06737-7

14. Finkelstein A, Cao X, Liao C, Schifitto G, Zhong J. Diffusion encoding methods in mri: perspectives and challenges. Investig Magn Reson Imaging. 2022;26:208–219.

15. Taoka T, Masutani Y, Kawai H, Nakane T, Matsuoka K, Yasuno F, Kishimoto T, Naganawa S. Evaluation of glymphatic system activity with the diffusion MR technique: diffusion tensor image analysis along the perivascular space (DTI-ALPS) in Alzheimer’s disease cases. Jpn J Radiol. 2017;35:172–178. doi: 10.1007/s11604-017-0617-z

16. Taoka T, Ito R, Nakamichi R, Kamagata K, Sakai M, Kawai H, Nakane T, Abe T, Ichikawa K, Kikuta J, et al. Reproducibility of diffusion tensor image analysis along the perivascular space (DTI-ALPS) for evaluating interstitial fluid diffusivity and glymphatic function: CHanges in Alps index on Multiple conditiON acquIsition eXperiment (CHAMONIX) study. Jpn J Radiol. 2022;40:147–158. doi: 10.1007/s11604-021-01187-5

17. Schartz D, Finkelstein A, Hoang N, Bender MT, Schifitto G, Zhong J. Diffusion-Weighted Imaging Reveals Impaired Glymphatic Clearance in Idiopathic Intracranial Hypertension. AJNR Am J Neuroradiol. 2024;45:149–154. doi: 10.3174/ajnr.A8088

18. Tian Y, Cai X, Zhou Y, Jin A, Wang S, Yang Y, Mei L, Jing J, Li S, Meng X. Impaired glymphatic system as evidenced by low diffusivity along perivascular spaces is associated with cerebral small vessel disease: a population-based study. Stroke and Vascular Neurology. 2023;8:e002191.

19. Sims JR, Gharai LR, Schaefer PW, Vangel M, Rosenthal ES, Lev MH, Schwamm LH. ABC/2 for rapid clinical estimate of infarct, perfusion, and mismatch volumes. Neurology. 2009;72:2104–2110. doi: 10.1212/WNL.0b013e3181aa5329

20. von Elm E, Altman DG, Egger M, Pocock SJ, Gotzsche PC, Vandenbroucke JP, Initiative S. The Strengthening the Reporting of Observational Studies in Epidemiology (STROBE) statement: guidelines for reporting observational studies. Lancet. 2007;370:1453–1457. doi: 10.1016/S0140-6736(07)61602-X

21. Schartz D, Finkelstein A, Bender M, Kessler A, Zhong J. Association of Extent of Transverse Sinus Stenosis With Cerebral Glymphatic Clearance in Patients With Idiopathic Intracranial Hypertension. Neurology. 2024;103:e209529. doi: 10.1212/WNL.0000000000209529

22. Nguchu BA, Zhao J, Wang Y, de Dieu Uwisengeyimana J, Wang X, Qiu B, Li H. Altered Glymphatic System in Middle-Aged cART-Treated Patients With HIV: A Diffusion Tensor Imaging Study. Front Neurol. 2022;13:819594. doi: 10.3389/fneur.2022.819594

23. Tournier JD, Smith R, Raffelt D, Tabbara R, Dhollander T, Pietsch M, Christiaens D, Jeurissen B, Yeh CH, Connelly A. MRtrix3: A fast, flexible and open software framework for medical image processing and visualisation. Neuroimage. 2019;202:116137. doi: 10.1016/j.neuroimage.2019.116137

24. Bhushan C, Haldar JP, Choi S, Joshi AA, Shattuck DW, Leahy RM. Co-registration and distortion correction of diffusion and anatomical images based on inverse contrast normalization. Neuroimage. 2015;115:269–280. doi: 10.1016/j.neuroimage.2015.03.050

25. Avants B, Tustison N, Song G. Advanced normalization tools (ANTS). Insight J. 2008;1–35. doi: 10.54294/uvnhin

26. Toh CH, Siow TY. Glymphatic Dysfunction in Patients With Ischemic Stroke. Front Aging Neurosci. 2021;13:756249. doi: 10.3389/fnagi.2021.756249

27. Qin Y, Li X, Qiao Y, Zou H, Qian Y, Li X, Zhu Y, Huo W, Wang L, Zhang M. DTI-ALPS: An MR biomarker for motor dysfunction in patients with subacute ischemic stroke. Front Neurosci. 2023;17:1132393. doi: 10.3389/fnins.2023.1132393

28. Zhu J, Mo J, Liu K, Chen Q, Li Z, He Y, Chang Y, Lin C, Yu M, Xu Y, et al. Glymphatic System Impairment Contributes to the Formation of Brain Edema After Ischemic Stroke. Stroke. 2024;55:1393–1404. doi: 10.1161/STROKEAHA.123.045941

29. Gaberel T, Gakuba C, Goulay R, Martinez De Lizarrondo S, Hanouz JL, Emery E, Touze E, Vivien D, Gauberti M. Impaired glymphatic perfusion after strokes revealed by contrast-enhanced MRI: a new target for fibrinolysis? Stroke. 2014;45:3092–3096. doi: 10.1161/STROKEAHA.114.006617

30. Liang D, Bhatta S, Gerzanich V, Simard JM. Cytotoxic edema: mechanisms of pathological cell swelling. Neurosurg Focus. 2007;22:E2. doi: 10.3171/foc.2007.22.5.3

31. Gu Y, Zhou C, Piao Z, Yuan H, Jiang H, Wei H, Zhou Y, Nan G, Ji X. Cerebral edema after ischemic stroke: Pathophysiology and underlying mechanisms. Front Neurosci. 2022;16:988283. doi: 10.3389/fnins.2022.988283

32. Meyer JS. Early adrenalectomy stimulates subsequent growth and development of the rat brain. Experimental neurology. 1983;82:432–446.

33. Winkler L, Blasig R, Breitkreuz-Korff O, Berndt P, Dithmer S, Helms HC, Puchkov D, Devraj K, Kaya M, Qin Z. Tight junctions in the blood–brain barrier promote edema formation and infarct size in stroke–ambivalent effects of sealing proteins. Journal of Cerebral Blood Flow & Metabolism. 2021;41:132–145.

34. Pirici I, Balsanu TA, Bogdan C, Margaritescu C, Divan T, Vitalie V, Mogoanta L, Pirici D, Carare RO, Muresanu DF. Inhibition of Aquaporin-4 Improves the Outcome of Ischaemic Stroke and Modulates Brain Paravascular Drainage Pathways. Int J Mol Sci. 2017;19. doi: 10.3390/ijms19010046

35. Sylvain NJ, Salman MM, Pushie MJ, Hou H, Meher V, Herlo R, Peeling L, Kelly ME. The effects of trifluoperazine on brain edema, aquaporin-4 expression and metabolic markers during the acute phase of stroke using photothrombotic mouse model. Biochim Biophys Acta Biomembr. 2021;1863:183573. doi: 10.1016/j.bbamem.2021.183573

36. Lv T, Zhao B, Hu Q, Zhang X. The Glymphatic System: A Novel Therapeutic Target for Stroke Treatment. Front Aging Neurosci. 2021;13:689098. doi: 10.3389/fnagi.2021.689098

37. Taoka T, Fukusumi A, Miyasaka T, Kawai H, Nakane T, Kichikawa K, Naganawa S. Structure of the Medullary Veins of the Cerebral Hemisphere and Related Disorders. Radiographics. 2017;37:281–297. doi: 10.1148/rg.2017160061

38. Taoka T, Ito R, Nakamichi R, Nakane T, Kawai H, Naganawa S. Diffusion tensor image analysis ALong the perivascular space (DTI-ALPS): revisiting the meaning and significance of the method. Magnetic Resonance in Medical Sciences. 2024:rev. 2023-0175.

39. Ringstad G. Glymphatic imaging: a critical look at the DTI-ALPS index. Neuroradiology. 2024;66:157–160.

40. Taoka T, Naganawa S. Glymphatic imaging using MRI. Journal of Magnetic Resonance Imaging. 2020;51:11–24.

41. Finkelstein AJ, Liao C, Cao X, Mani M, Schifitto G, Zhong J. High-fidelity intravoxel incoherent motion parameter mapping using locally low-rank and subspace modeling. Neuroimage. 2024;292:120601. doi: 10.1016/j.neuroimage.2024.120601

